# Evaluating The Feasibility and Effectiveness of a Capacity-Building Model to Nurture Junior Independent Clinical Research Investigators in Uganda

**DOI:** 10.1101/2025.10.10.25337759

**Authors:** Aidah Nanvuma, Joseph Musaazi, Adelline Twimukye, Vivian Nakate, Agnes Kiragga, Miriam Laker Oketta, Christine Sekaggya-Wiltshire, Yukari. C. Manabe, Barbara Castelnuovo

## Abstract

**Background:** Research capacity-building initiatives remain crucial to achieving Sustainable Development Goal 3 on health and well-being, especially in LMICs. We aimed to evaluate the effectiveness of the Infectious Disease Institute’s (IDI) capacity-building model to nurture junior independent clinical research investigators in Uganda.

**Methods:** From 13^th^ July 2021 to 06^th^ February 2023, we conducted a trend analysis study using a mixed methods approach to assess the extent to which the Capacity-Building Model was effective, feasible, and acceptable. For quantitative research, we conducted an online survey with 80 trainees (Masters, PhD, and Postdoctoral fellows), comprising 20 alumni and 60 current trainees, to explore their experiences and perceptions as former and current scholars of the Capacity Building Unit (CBU) at IDI. For qualitative research, we purposively selected 20 trainees to participate in the in-depth interviews.

**Results:** Participants reported that the capacity-building model had a beneficial impact on their career progression, with 90% expressing a willingness to recommend it to others. The overall scientific benefit reported was 48.7%; this was significantly higher among continuing scholars compared to alumni (56.7% vs 25.0%, respectively, p-value = 0.046). Additionally, 85% achieved their career goals, and 65% said it expanded their employment opportunities. Additionally, 85% achieved their career goals, and 65% said it expanded their employment opportunities. Qualitative findings highlighted its significant positive impact on research training and professional development. Participants praised the model’s emphasis on mentorship, with both scientific and non-scientific support proving crucial in guiding junior researchers through technical challenges, manuscript writing, and career planning. Soft skills training, dissemination platforms, and networking opportunities further contributed to scholars’ academic growth. Trainees benefited from robust institutional support, including access to research infrastructure, grantsmanship assistance, and administrative systems. However, challenges such as limited research funding, slow procurement processes, and supervisory delays hindered progress. The COVID-19 pandemic also disrupted mentorship and training. Participants recommended improvements in mentorship coordination, procurement efficiency, broader model visibility, and expansion to other universities and disciplines to enhance its effectiveness and sustainability.

**Conclusion:** Overall, the acceptability of the Capacity-Building Model was high among scholars; however, minor administrative challenges need to be addressed to enhance learning further. We recommend tailored, relevant scholarly programs to meet the evolving needs of emerging scientists and foster scholarly growth and research innovation in academic institutions, enabling them to tackle local public health challenges. Future research is required to assess the cost of capacity building and its sustainability.

## INTRODUCTION

### Background

Research capacity building remains crucial for enhancing health outcomes in resource-constrained settings (1). This entails intentionally developing sustainable individual and institutional (2, 3) research capacity-building skills, structures, and resources to achieve Sustainable Development Goals (SDG), particularly SDG 3 on Good Health and well-being (4). However, formal research training in institutions of higher learning remains limited (5).

Training is usually complemented by mentorship. Mentorship involves providing guidance, knowledge, and support from an experienced individual, referred to as a mentor, to a less experienced individual, known as a mentee, to help them navigate a career pathway (6, 7). The primary objective of mentorship relationships is to enable the mentor and mentee to acquire skills in the proper conduct of research, dissemination of research results, and knowledge management and utilization. Prior studies report two common forms of mentorship: peer-to-peer and senior-to-junior mentorship (8). With peer-to-peer mentorship, students or colleagues with similar experiences, ages, and levels of power engage in reciprocity by offering psychosocial support and facilitating learning exchanges (9, 10) with one another. The senior-to-junior format may involve an experienced faculty member mentoring a less experienced member to transfer skill sets and knowledge.

Mentorship remains fundamental in nurturing the next generation of researchers, as valuable knowledge and expertise are passed down, thereby contributing to the growth of research capacity. Moreover, developing professional networks facilitates collaboration and knowledge sharing among researchers locally and globally (9). These networks provide valuable opportunities for researchers to exchange ideas, collaborate on projects, and access resources and expertise beyond their immediate environments (10).

Despite notable progress in health research capacity development in LMICs, there remains substantial room for improvement (10). Few programs focus on investing in, or implementing systemic or institutional-level approaches to, capacity development (11–13). We, therefore, reflected on the operational aspects of a Sustainable Research Capacity Building Model used to nurture credible researchers at the Infectious Diseases Institute, Uganda (14), to solicit research trainee experience and appraisal of our systems and infrastructure towards nurturing independent research investigators.

## METHODOLOGY

### Study design

From 13^th^ July 2021 to 06^th^ February 2023, we conducted a cross-sectional study using a mixed-methods approach to evaluate the effectiveness, feasibility, and acceptability of the Capacity-Building Model used at IDI. For the quantitative component, a structured online survey was administered to Master’s, PhD, and postdoctoral trainees, including both alumni and current scholars, to explore their experiences and perceptions of the Capacity Building Unit (CBU) programs. Additionally, we conducted 20 in-depth interviews with participants across all training levels to gain a deeper understanding of their experiences. Alumni were defined as trainees who had published their research in a peer-reviewed journal, while current scholars were those who had not yet published, regardless of their time in the program. Data collection took longer than expected due to the COVID-19 interruption in 2021.

### Study setting

#### Infectious Diseases Institute

The IDI was established in 2002 under the College of Health Sciences at Makerere University. It is a Ugandan not-for-profit organization dedicated to strengthening Africa’s health systems, with a focus on infectious diseases, through research and capacity development. IDI is located opposite the new Mulago Hospital complex and near Makerere University’s main campus. IDI comprises eight departments: Research, Prevention Care and Treatment, Health System Strengthening, Training, Global Health Security, and Laboratory Services. It employs over 1,000 staff members. IDI provides care and treatment to more than 209,000 people living with HIV across urban and rural Uganda, both directly through its large clinic and in partnership with government and non-government health facilities. The Institute also supports long-term outreach programs aimed at building capacity nationwide, with over 70,000 healthcare workers trained to ensure responsive and equitable health service delivery. The IDI Research Department has overseen over 200 studies, with 70 active projects currently underway, including clinical trials, observational studies, diagnostic and implementation studies, as well as student research at the master’s and Ph.D. levels. To date, over 1,300 researchers have published more than 1,300 peer-reviewed journal articles (15). The CBU operates under the leadership of the Head of the Research Department at IDI.

### Description of the Capacity Building Model at IDI

The Infectious Diseases Institute established a Capacity Building Unit specifically designed for young investigators. This initiative aims to improve their research skills and competencies, thereby addressing the training gap and fostering a more robust research environment. The Capacity-Building initiative aims to enhance research capacity by providing various training opportunities, mentorship, and research fellowships (Including Postdoctoral, Ph.D., and Master’s programs) to supplement research-related courses. These training courses are offered onsite, offsite, and online, including short courses such as Epidemiology, data analysis, and Good Clinical Practice, with a focus on developing well-rounded, independent researchers at the Infectious Diseases Institute (IDI) and collaborating institutions, which are both local and international. The Capacity Building Model (Fig.1) at IDI is based on two principles: 1) Senior to junior and 2) Peer mentorship (16). While most IDI scholars were previously mentored by international experts, in recent years, mentorship has shifted gradually to local senior scientists.

**Fig. 1.**
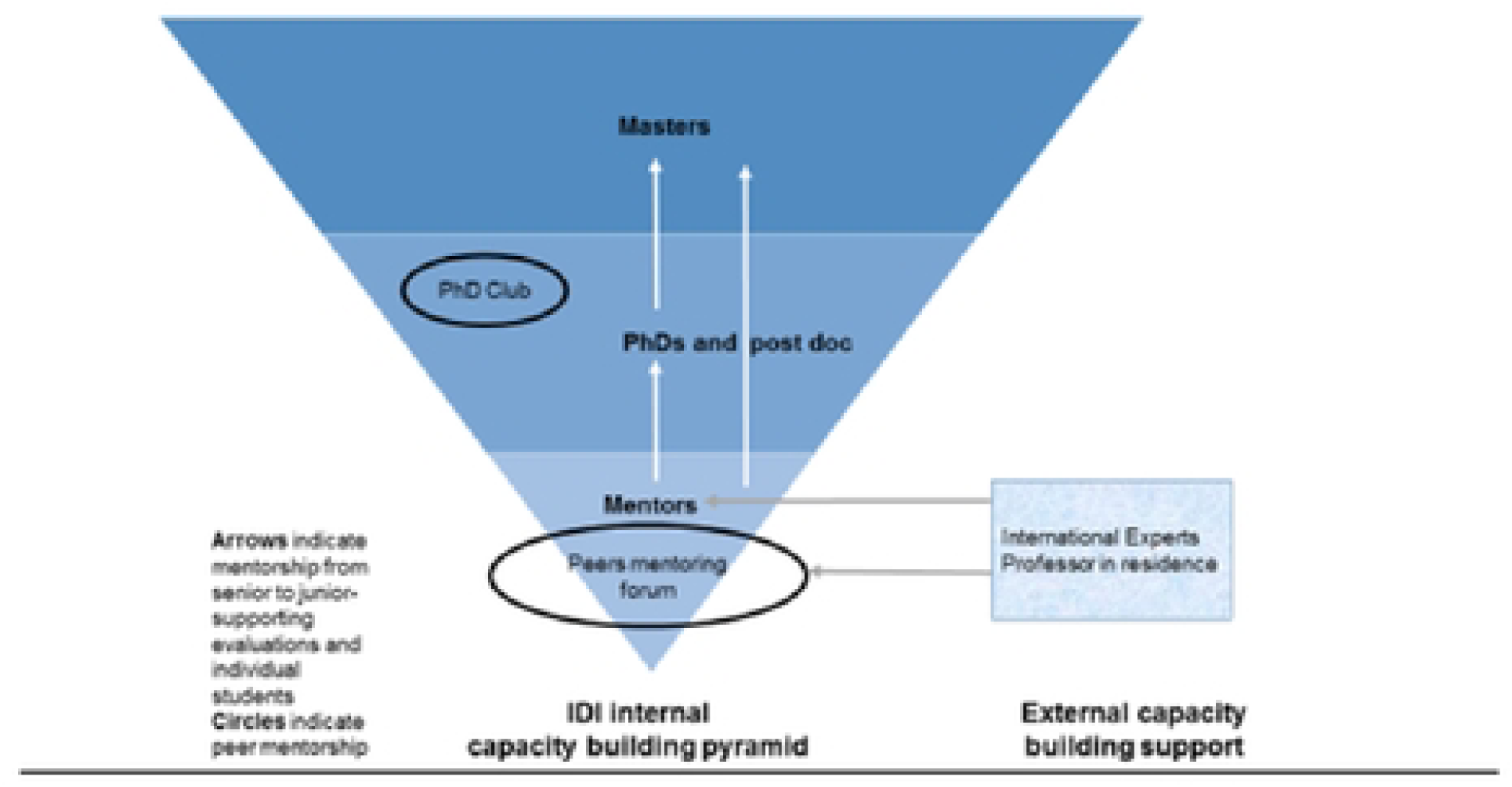
Shows the sustainable capacity-building model at IDI.

The local senior scientists, in turn, are all IDI alumni who had international mentors. Some degree of responsibility has also been shifted to Ph.D. scholars, who were assigned as co-mentors to master scholars to implement the concept of sustainable research capacity building. The other component of capacity building is peer-to-peer mentorship. We have successfully established a Ph.D. club, an emerging scientists’ group (mostly study coordinators), a social science group, a research fellows group, and a senior scientists (mentors) group, where members mentor each other by presenting their proposals or results, discussing journal articles, or teaching each other advanced research methodologies and soft skills. International experts, in this model, still play a crucial role in advising the senior scientists (on their research or how to mentor the junior scientists best on focused areas where the local expertise or technology is still lacking, for example, laboratory science. Of note, due to its richness in local scientific expertise, rigorous grants management, and variety in the exposure to infectious diseases, IDI is a training site for Makerere University-both medical students and masters in internal medicine students for rotations and has become a popular site for the placement of international scholars from existing collaborations (e.g., the University of Minnesota, University of Zurich, University of Turin) and an affiliated site for some competitive US university capacity building grants (e.g., Fogarty GloCal Health Fellowships)

### Participant selection and sampling

Study participants for the quantitative survey were approached by email to provide consent to participate in the interviews. We interviewed 80 trainees under the capacity building unit, comprising alumni and continuing trainees. We included all trainees affiliated with IDI CBU since its inception who were available and provided consent to participate in the study. For qualitative data, we conducted 20 in-depth interviews, continuing with trainees until saturation was achieved, based on analysis of detailed notes and debrief summaries. The participants’ lists were generated from CBU records with contact details such as email and telephone numbers.

### Data collection

We conducted an online survey using KoBo Toolbox. An email with a link to the online Informed Consent Form and the survey questionnaire was sent to all participants to solicit their participation. To increase the response rate from study participants, follow-up emails and telephone call reminders were sent to participants who had not completed the survey within 1 to 2 weeks after sending the online questionnaire response invitation. Data were collected using a pre-coded questionnaire in the English language. The questionnaire consisted of prompts on baseline demographic characteristics, including age, gender, and date of birth. Additionally, responses are sought on scholars’ experiences (pre-, during, and post-) interacting with the Unit’s programs and activities. Participants were asked to rank unit activities (Scientific and non-scientific support in Figure 2, provided as described above) that were considered most beneficial regarding the expected scholarly outputs and brief explanations. Data on challenges encountered by scholars while under the Unit, programmatic gaps, and suggested areas for improvement were assessed, along with post-training attitudes and beliefs. A Likert scale of one to five will be used for the survey questions to rank the contribution of unit support activities to scholarly outputs. Where; 1=Strongly Disagree, 2=Disagree, 3= Neutral (Undecided), 4=Agree, and 5 =Strongly Agree. The Likert scale data will be categorized into three groups: strongly disagree’ and “disagree” (Low), “neutral” - medium, and “agree” and “strongly disagree (high), and after that, handled as a categorical variable, and any differences between the categorical variables will be tested using chi-square tests. Participants were only able to submit their responses after completion. For qualitative data, we conducted in-depth interviews with Master’s scholars, Ph.D. scholars, and postdoctoral scholars. We collected data using a topic guide to assess participant experiences, challenges, gaps, and recommendations for unit programs and activities.

**Fig. 2.**
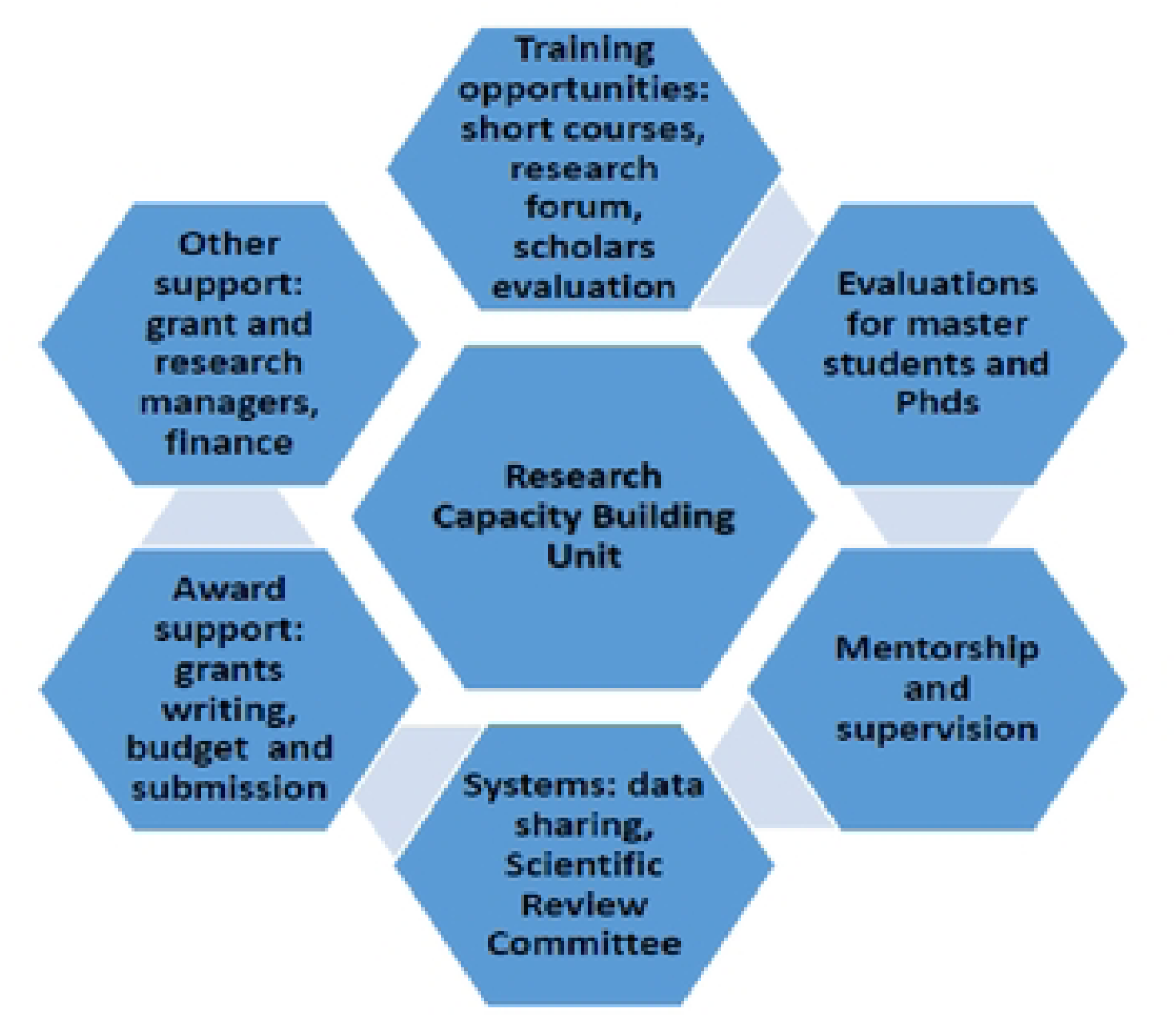
Scientific and non-scientific support provided by the Capacity Building Unit to research trainees at IDI.

Additionally, we assessed scholar attitudes and beliefs post-training and current engagements. Interviews were conducted in English, audio recorded, and lasted between 30 to 45 minutes. Participants provided a written informed consent form before participation. The data collection and analysis period lasted six months.

### Data management and analysis

Quantitative data were extracted from Google Forms and exported to Microsoft Excel 2016 for cleaning and coding. The cleaned dataset was exported to STATA (v15.1) for analysis, categorized into alumni and continuing scholars, and summarized using frequencies and percentages. An inductive thematic approach was used for data analysis, focusing on qualitative data derived from a topic guide. Three themes were identified from the questionnaire analysis, including the effectiveness and approaches of the capacity-building model, challenges or gaps in the model, and recommendations.

Researchers reviewed transcribed in-depth interviews to familiarize themselves with the data. A joint coding framework was developed from five transcripts representing 25% of the total to enhance consistency and transparency in coding. Two coders (AT and JB) performed open coding. Initial codes were refined through consensus and consultation with the Principal Investigator, and reliability was ensured by cross-checking among coders. The transcripts were organized and imported into NVivo version 12 software for open coding, which led to the categorization of codes and the identification of themes. A final merged codebook synthesized key findings, and illustrative quotations were selected to represent each theme in the results.

## RESULTS

### Social demographics characteristics of the study participants

A total of 80 participants were enrolled, comprising 20 alumni and 60 continuing research scholars at the IDI Capacity Building Unit at the Infectious Diseases Institute, as shown in Table 1. The median age of alumni was 35 years (IQR: 30.5-42.0) and 34 years for continuing scholars (IQR: 29.5-38.0). Most Trainees of both groups had completed or were pursuing a Master’s degree (alumni: 85.0%, continuing scholars: 65.0%). There was a slightly higher representation of males in both groups: alumni (65.0%) and continuing scholars (66.7%).

**Table 1:** Social Demographics Participant characteristics.

| **Variable** | **Alumni scholars**<br>**(20, 25%)**<br>**n (%) or median**<br>**(IQR)** | **Continuing scholars**<br>**(60, 75%)**<br>**n (%) or median**<br>**(IQR)** | **Total**<br>**(80, 100%)**<br>**n (%) or median**<br>**(IQR)** |
| --- | --- | --- | --- |
| **Age in years**<br>(Median [IQR]) | 35 (30.5 - 42.0) | 34 (29.5 - 38.0) | 34 (30-38) |
| **Education level completed or pursuing.** |  |  |  |
| Masters | 17 (85.0) | 39 (65.0) | 56 (70.0) |
| PHD | 2 (10.0) | 13 (21.7) | 15 (18.8) |
| Post Doc | 1 (5.0) | 8 (13.3) | 9 (11.3) |
| **Sex** |  |  |  |
| Male | 13 (65.0) | 40 (66.7) | 53 (66.3) |
| Female | 7 (35.0) | 20 (33.3) | 27 (33.8) |
| **Years of experience After First Degree** |  |  |  |
| <=5 years | 5 (25.0) | 22 (36.7) | 27 (33.8) |
| >5 years | 15 (75.0) | 38 (63.3) | 53 (66.3) |

**Table 2.**
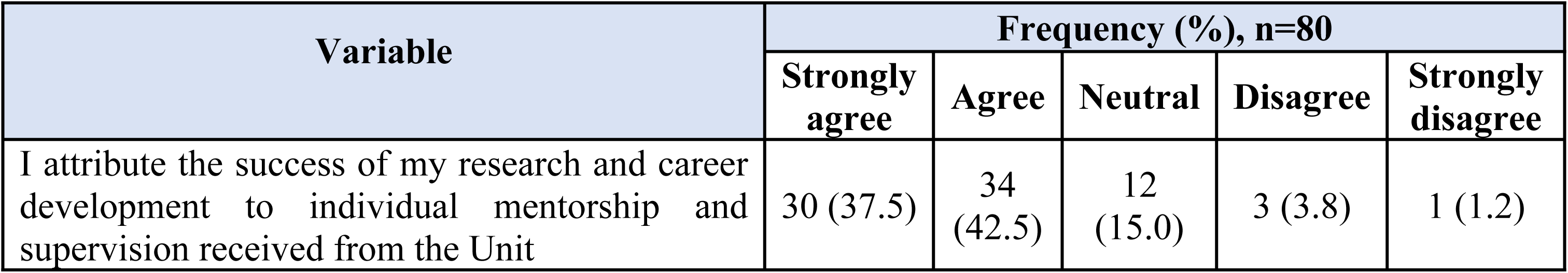
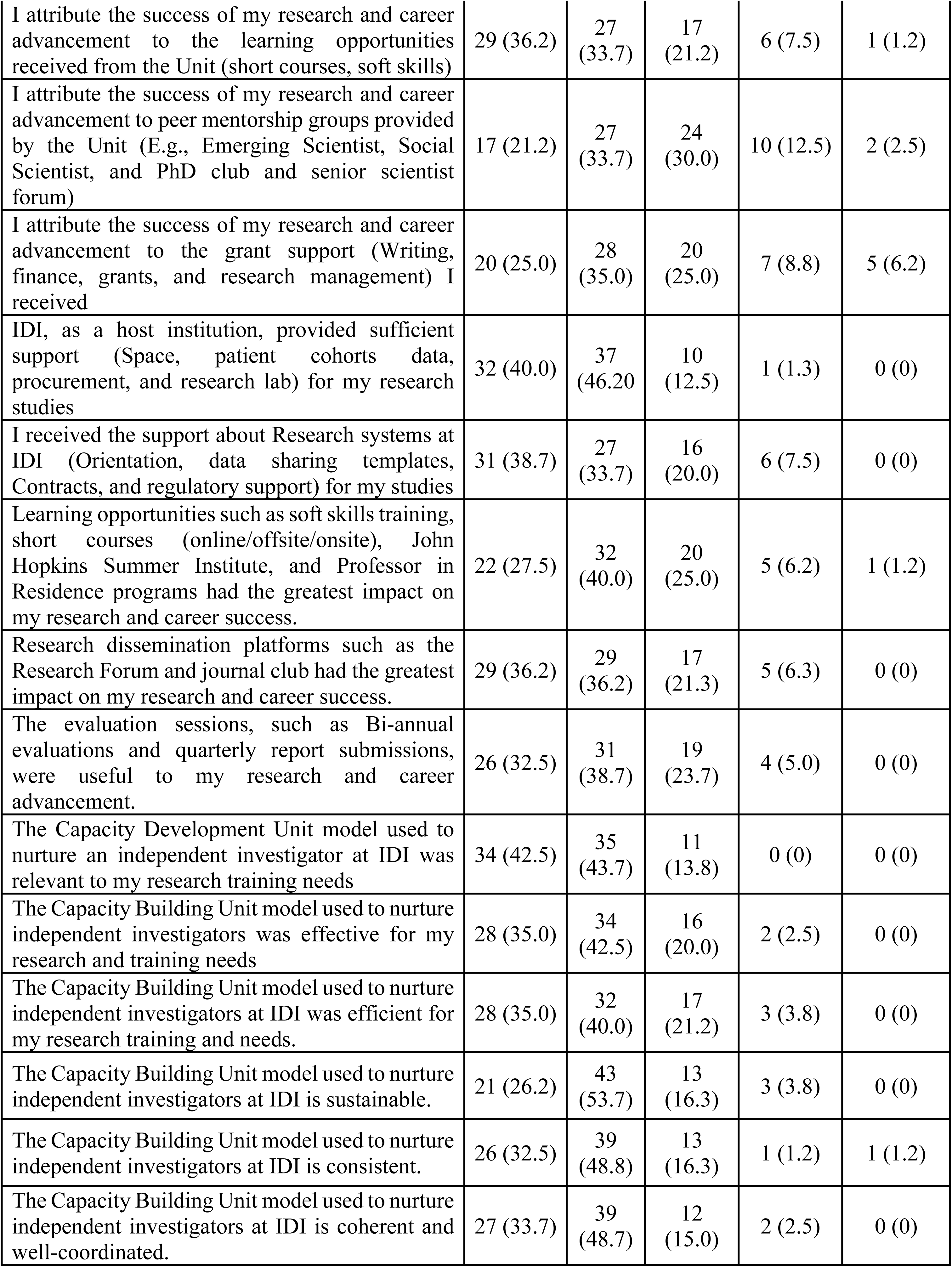
Scientific and Non-Scientific Support Provided by the Capacity Building Unit to IDI Alumni and Current Trainees.

### Feasibility and Effectiveness of The Capacity-Building Model

#### Capacity Building Model Effectiveness

Participants reported that the capacity-building model had a beneficial impact on their career progression, with 90% expressing a willingness to recommend it to others. The overall scientific benefit reported was 48.7%; this was significantly higher among continuing scholars compared to alumni (56.7% vs 25.0%, respectively, p-value = 0.046; Table 3). Additionally, 85% achieved their career goals, and 65% said it expanded their employment opportunities. Additionally, 85% achieved their career goals, and 65% reported that it expanded their employment opportunities (Fig. 3).

**Fig. 3.**
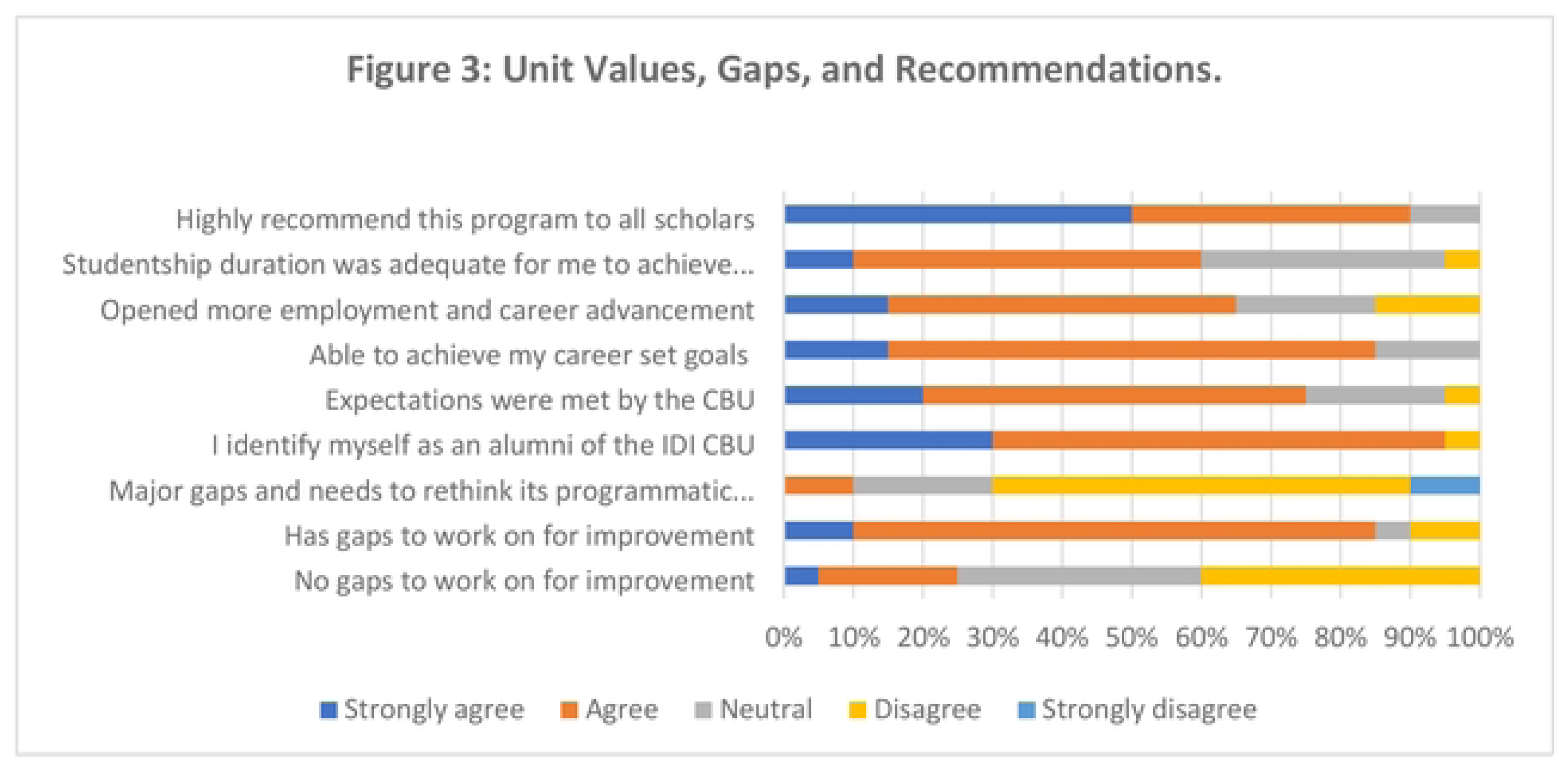
Unit Values, Gaps, and Recommendations.

### Approaches to Capacity Building Model

Participants described several capacity-building approaches that significantly supported their academic growth, with mentorship and support supervision emerging as central pillars. Respondents emphasized that individual scientific and non-scientific mentorship offered critical technical guidance, regular progress evaluation, and constructive feedback on research manuscripts. This mentorship was seen as instrumental in helping them acquire the skills and tools needed for academic and professional advancement. Many scholars have highlighted the value of a supportive supervision model that responds to their needs in real-time, facilitating faster progress in their research work. They expressed appreciation for mentors who possessed deep knowledge and leadership experience, noting these qualities as key to meaningful guidance. In addition to technical mentorship, participants recognized the importance of soft skills training, including writing, communication, and critical thinking, which they felt contributed to their overall development as well-rounded researchers. They also reported that platforms for research dissemination, such as weekly forums and journal clubs, exposed them to diverse perspectives and opened doors for collaboration. Support from the research office, particularly in terms of ethical review processes and grantsmanship, was also noted as crucial in helping them navigate complex academic environments. Networking events and targeted workshops were credited with promoting collaboration and the exchange of ideas. At the same time, the strong research infrastructure at the IDI was repeatedly mentioned as a foundational resource that enabled effective implementation of research activities.

Overall, more than 60(68.26%) of the trainees attributed their research and career development success to scientific support. In comparison, over 70% demonstrated that non-scientific support played a significant role in their research and career advancement. Mentors provided scholars with technical guidance for their research work, conducted progress evaluations, and offered constructive criticism on the manuscripts they reviewed. Through mentorship, participants were equipped with the necessary tools, skills, and support to thrive in their academic, research, and professional endeavors. The supervision design enabled scholars to progress faster because supervisors were supportive and responsive to time needs, for instance, by honoring appointments. The qualities of mentors desired by mentees were a rich Knowledge base, leadership, and research experience. Scholars or students identified their mentors as experts in the field of their pursuit.

> *“I got one of the best mentors within the unit, I do not have international mentors attached to me, but I am within a network where I benefit from a number of international mentors where the opportunities are provided…indirectly I think I do benefit from that kind of advice or information or technical guidance from international mentors and also across the country I think the platforms have helped us to benefit from even others that are not within the IDI setting”* -- IDI Male 1, Post-doc.

### Learning opportunities Soft skills

Almost all respondents agreed that they had benefited from soft skills training, including writing manuscripts and grants, communication, presentation, and critical thinking skills. Investing in soft skills development was beneficial for the holistic growth of junior researchers, contributing to their personal and professional success and empowerment, thereby facilitating progress. Soft skills were targeted for scholars, fellows, and staff. Some scholars reported receiving training in soft skills, including time management and mentorship.

> *“They [Soft skills] were great; they helped me improve my writing skills. I remember from them I discovered that presenting my data using pie charts, graphs was the best way, so they helped me to present my work better”* -- IDI Male 2, Masters.

### Research findings dissemination

Research findings dissemination platforms, such as research forums held weekly on Thursdays and journal clubs on Fridays, conducted either physically or via Zoom to critique papers, research work, and concepts, were found beneficial. They served as valuable resources for junior researchers, supporting their exposure to diverse perspectives, career growth, and collaboration with the research community. All staff, scholars, and collaborators could access the research forum, which enabled them to learn from other researchers and present their research concepts and ideas. The outcome of the research forum was receiving feedback on concepts presented and exchanging ideas that helped the scholars improve their ideas.

> *“…the research forums; the Thursday research forums were beneficial in way that, one they are like dissemination let say some has had their studies we are able to learn of their findings and in the process, you are taking through the whole process of…the whole process of research and updates…”* -- IDI Male 6, Masters.

> *“we have been having the Friday soft skill sessions one In statistics, as far as how to make a power point presentation by Doctor…Uhm then we have had mentors of who just come to share their experience the we’ve had the journal clerk where one us presents an article, I have also been one of the beneficiaries because I have also presented at the journal club and then yes the experience in its self has a way of building capacity…has a way of building capacity to the scholars”*--IDI Male 6, Masters.

### Support structures from the research office

The availability of support structures from the research office, including Internal Scientific review systems and collaborations, ethical bodies, and administrative clearances. These initiatives helped junior scholars navigate the complexities of academic research, enhance their skills, and foster professional development.

> *“These other systems of course the ethical bodies and all that, I had to present my work, again, at that proposal development stage, I had to present, that time to the IDI scientific review committee and from there actually I got more, I got to know some insights. So actually, each stage of, each stage where you present your work, there is something new that comes up. There is something new that is added, there is something new that is subtracted. Or there is something that is subtracted and there is something new that is added. So, these are really like good systems in place”* -- IDI Male 2, Ph.D.

### Grantsmanship support

Grantsmanship support was helpful throughout the scholars’ study journey. Most scholars said they were currently working on other grants. The Grants and Finance Department offered IDI staff, fellows, and scholars grant-writing support. The Research Office provides support in writing, finance, grants, and research.

> *“… I’ve been able to get all of those financial privileges. And I got-I was able to get a stipend as I told you. Plus, that fifty per cent 50 % time off which gave me the ability to do some other things to write the grants. Because grant writing especially as a person who is new, am a-I should say am an early career research I needed the time to think through these concepts so I got all that. So, to me, I’ve really gotten all the privileges”* -- IDI Female 2, Post-doc.

### Networking opportunities and workshops

Additionally, the office facilitated networking opportunities and workshops that promoted collaboration and knowledge sharing, thereby contributing to the growth and success of junior researchers in their academic careers. Scholars benefited from networking with experienced scientists or experts from different supportive fields. These are either field-based recommendations or sometimes events at the Institute. Scholars also established global or international networks through networking with experts and attending conferences.

> *“Then of course the networking, always referring you to someone who is going to help you, someone who has been there, someone who has experience in that matter and that’s it. It has really opened up my network and even this short time that I have been there. And then of course the help that I get from the rest of the scholars, because they are always giving little bits of advice. You do this, how to avoid this pitfall, you know things like that which are also very useful then of course the access to all the other things”* -- IDI Female 1, Ph.D.

### Good infrastructure and ecosystem are available at IDI

Good infrastructure and ecosystem are available at IDI, which enhances learning. IDI was described as a world-class research environment with well-established research systems that enhanced learning for junior researchers. Participants stated that they accessed institutional resources, such as computers or laptops, the Internet, printing services, and access to scholarly articles through the library at no cost, as well as obtained statistical help from qualified statisticians. These resources enabled them to gather and analyze data, collaborate with peers, and produce high-quality written work. Reliable Internet access enabled efficient communication and access to online databases and research materials. At the same time, printing facilities ensured that they could produce physical copies of their work for presentations or submissions, thereby enhancing productivity and facilitating the research process for junior scholars.

> *“They gave me someone to help me, they have a system, when you’re analysing your data, a statistician will help you to confirm whether what you’re doing is right. I think the only difference is for me I had done bio statistics before the IDI session was engaged, I had actually analysed my data, through it was my first time to analyses a lot of data like going through the process and I really got feedback from the statistician I worked with and she motivated me to continue doing a number of things”* -- IDI Male 2, Masters.

> *“I think at IDI the research systems are really very well established, the eco system of research in Uganda is top notch…, but that is what I think happened but for the eco systems of research IDI is like a university of its own, they have all the resources and infrastructure for a person to practice research. But in my own view it wasn’t clear to people because even when I inquired, they said that you know we are going to get you a slot as a scientist as you know you must have to first write your own grants, that sort of things…”*--IDI Male 3, Post-doc.

### Challenges or gaps in the capacity-building model

This section outlines several significant challenges faced by scholars within the Capacity Building Unit (CBU), with a primary focus on inadequate research funding. Scholars reported financial constraints as a major obstacle, with limited access to sufficient funding impacting their ability to conduct research, access necessary resources, and secure essential materials. This lack of funding stifled their professional development and collaboration opportunities, increasing stress and uncertainty regarding their career prospects and potentially leading to decreased innovation and talent loss within the research community. Additionally, the text highlights other challenges, such as procurement delays, supervisory delays, and interruptions caused by the COVID-19 pandemic, all of which further hindered the research progress and productivity of emerging scientists.

### Programmatic or administrative challenges

Figure 3 shows that 10% of the participants stated that the Unit had programmatic or administrative challenges among notable challenges experienced by continuing scholars before joining the CBU, which decreased during their time at the CBU, with varying perceptions at different stages: 15.0% strongly agreed, 41.7% agreed before joining, decreasing to 8.3% strongly agreed and 18.3% agreed during the CBU tenure. Regarding alumni respondents during the pre-training phase, when scholars were prepared to join the CBU, 30% agreed that they encountered challenges, 15% were neutral, and 40% disagreed. Similarly, during the in-training phase, where scholars actively engaged in training activities, 30% agreed they faced challenges, 20% were neutral, and 40% disagreed. Programmatic or administrative challenges reported in the qualitative data included inadequate research funding, procurement delays, supervisory delays, and COVID-19-related interruptions.

### Inadequate research funding

Scholars reported financial constraints as a major challenge encountered while under CBU, as they had limited access to sufficient funding. Some respondents did not have enough funds to conduct their research, which was not anticipated during their engagement with the program. The stipend provided was limited, especially for scholars who lacked other sources of income. Insufficient funding created barriers that stifled the growth and contributions of emerging scientists. It limited their ability to conduct experiments, access necessary resources, and secure essential materials. This financial constraint hindered their professional development, reduced opportunities for collaboration, and diminished their chances of publishing impactful research. Consequently, junior researchers faced increased stress and uncertainty regarding their career prospects, which could potentially lead to a decrease in innovation and a loss of talent within the research community.

> *“I think my other challenge that I faced was in terms of research funding…I experienced that; my research included me recruiting patients and then doing all doing culture studies…and the Unit did not have additional funding”* -- IDI Female 1, Masters.

### Procurement delays

Procurement bureaucracy was reported as a major challenge to CBU. The slow procurement process resulted in delays in purchasing research equipment, such as laptops, to support the analysis process. It involved a lot of back and forth from different personnel, hence the failure of the process. Procurement delays had a detrimental impact on emerging scientists, hindering their research progress and productivity. When essential materials, equipment, or resources were not delivered on time, it resulted in interruptions to experiments and project timelines. This resulted in missed deadlines for grant applications, publications, and presentations, which are critical for career advancement. Additionally, such delays increased frustration and stress, potentially impacting the mental well-being of emerging scientists.

> *“Actually, I wanted to procure a laptop because it was going to be a center of my research; Well, its quite a lengthy process from my experience…”* -- IDI Male 3, Masters

### Supervisory delays

Students were significantly affected by delays in receiving feedback. These delays were mainly caused by external supervisors, but some were also due to busy supervisor schedules, which contributed to the delays and hindered their research progress. Some also noted that having many supervisors was associated with discrepancies in agreement, which would lead to numerous back-and-forths without making progress. They pointed out that with many supervisors, they were more likely to have varying opinions on concepts. When supervisor guidance and feedback were not provided in a timely manner, it slowed down the research process, resulting in prolonged project timelines. This was detrimental for junior researchers who rely on their supervisors for critical insights and direction, especially when navigating complex research challenges. Additionally, delays in supervision led to a lack of clarity regarding project goals and expectations, which may result in confusion and misalignment in research efforts. This uncertainty hindered creativity and innovation, as emerging scientists may hesitate to pursue new ideas without clear approval or support.

> *“I have five functional supervisors. Although if you look at my official documents it will say three. And it is very difficult to get all five of them to agree or give you feedback maybe at the same time. For example, as I have told you, I’ve just sent out a new version of my concept and so far, I have only received comments from one person and yet each of them has something to contribute to my concept and how I am going to move forward. So, I find that it is very difficult to move forward when some of them have not given me their comments and their advice and things like that. So, I find that that is slowing me down a little bit because I am always having to nudge and to wait”* -- IDI Female 1, Ph.D.

### COVID-19 interruptions

COVID-19 is reported to have disrupted mentor schedules by restricting the travel of some mentors who had planned to fly in and supervise students or mentees who needed training. It also disrupted physical interactions, yet some students still felt they needed physical sessions to gain practical exposure at that time for their respective courses. Junior scientists faced disruptions in their research activities due to the COVID-19 pandemic, laboratory closures, limited access to resources, and the cancellation of conferences and networking opportunities.

This led to delays in data collection and hindered progress on projects. Additionally, the shift to remote work created difficulties in collaboration and mentorship, which are crucial for professional development. Junior scientists also experienced increased stress and uncertainty regarding job security and career advancement as funding opportunities became more competitive and uncertain.

> *“the situation with COVID-19, has brought many changes. I mean, it has shifted my original plan. The scheduled work activities. I will give an example; there was a training that was scheduled for September in Johns Hopkins University. But it was cancelled because I was vaccinated with AstraZeneca, which is not recognized by the CDC. So according to the CDC policy on vaccination, it is like I am not vaccinated, so I cannot fly out of the country until maybe I get Johnson & Johnson or Moderna vaccines”* -- IDI Male 4, Ph.D.

### Recommendations from participants

This section emphasized the importance of publicizing the capacity-building model to enhance stakeholder awareness, facilitate knowledge sharing, attract funding, encourage collaboration, and establish standardized practices. It highlights that information about the model can be disseminated through advertisements, online platforms, and research events, allowing interested parties to benefit. Participants express the need for clearer communication regarding the model’s operations, particularly concerning diverse fields such as nursing and informatics. This suggests that better visibility could aid in orientation and understanding of the program. Figure 3 shows the willingness to recommend the capacity-building model.

### Publicization of the capacity-building model

Publicization of the capacity-building model through advertisement and information sharing to different institutions and the public raises awareness among stakeholders, facilitates knowledge sharing of best practices, attracts funding and resources, encourages collaboration among organizations, and promotes the establishment of standardized practices on how it operates, which is shared with the public through the Internet and physical research days. Publicizing the model will enable other interested parties to benefit from it.

> *“If we can open it up, people can see it divergently. Like for me, I’m from nursing, but I am thinking in line with informatics. So, I did not see it very clear whether this orientation because it is more of medicine…IDI to help, maybe to help a colleague, if we have information displayed like on a website and we can know okay this year the capacity building unit has this division, this discipline it becomes easier.* --- IDI Male 7, Masters.

### Scaling up of capacity building model

Scaling up the capacity-building model enhances its effectiveness and reach, resulting in more substantial and sustainable development outcomes. This can be done by building the capacity of students in other universities, not only in Makerere but also in the core research or medical team at the local Institution.

> *“The other thing I thought about would be, IDI expanding, so IDI being like— expanding this Capacity Building Programme beyond Makerere University. I don’t know the politics that would be involved but for instance IDI trying to train some faculty members from other Universities from say Gulu university, Busitema university inaudible segment] university because I understand IDI has some research programmes in Arua, I think in Mbale. So, if they train some individuals form such institutions then it becomes easier for them to undertake their programmes in such areas. Because I think, according to my knowledge it seems currently the Capacity Building Programme is emphasized in Makerere”* -- IDI Male 2, Post-doc.

Identify promising students from medical schools who are keen to join CBU. IDI can put out actual calls for research ideas to attract more researchers and even send them abroad.

> *“So far, the recruitment of scientists at IDI is mostly from the pool of people on the Mulago hill. But there are many research IDIs out there, so I was thinking maybe IDI can put up an actual call for research ideas because there are students who are truly promising if they can be identified from the medical schools, attached.”* -- IDI Male 3, Post-doc.

### Strengthen a multi-disciplinary approach

The adoption of a multi-disciplinary approach, which includes diverse research teams and topics, brings together varied expertise and viewpoints, leading to more comprehensive solutions and collaboration across disciplines. A multi-disciplinary approach could effectively engage with communicable diseases and involve people from policy programs, business fields, and basic research.

> *“I think at that level I will talk more on policy engagements since I am not involved in the patient management. I think an area where I think we need an improvement is if we can open more platforms where we can help researchers to engage policy makers that would be great. I think if we can have policy makers at all stages of research works from the time you identify a particular research area and develop proposals and get results and disseminate”* -- IDI Male 1, Post-doc.

### Strengthen mentorship coordination

The need for the Institution to strengthen mentorship coordination between mentees and mentors was suggested by participants to increase consensus on topics and proposals, thereby reducing students’ frustration when their supervisors from different units disagree on some agreed-upon course of action for protocol, leading to wasted time and energy. Strategies for Strengthening Mentorship Coordination for Junior Researchers include establishing formal mentorship programs, such as pairing junior researchers with experienced mentors based on interests and goals, with clear guidelines. Facilitate ongoing communication to discuss progress and challenges and offer timely feedback. Promote support among junior researchers through peer mentoring and strengthen peer group leadership and enrollment. Two scholars said they were not members of any peer groups.

> *“…at team level or group level, I think you know each team or group level I think you know each team or group leader who understands the team, and if we have groups, you really need to have a leader who understands the team and communicates the vision of the team or group clearly and people but in and I think this will help people to propel the team to achieve that common goal in a way it also improves personal like achievements so for groups and teams I would basically look at focusing more on the team leaders or group leaders and group leaders taking more time to understand the team members and supporting them”* -- IDI Male 1, Post-doc.

### Improvement of the procurement process

Improvement of the procurement process by providing comprehensive training on procurement principles, tools, and best practices. This can include workshops, online courses, or mentorship sessions that cover key topics such as supplier selection, contract negotiation, and compliance. Develop and document standardized procurement processes and templates. This can help the junior researchers understand the steps involved and ensure consistency in their work. Introduce procurement software or tools that can streamline the process, making it easier for the researchers to manage tasks such as sourcing, tracking orders, and analyzing supplier performance.

> *“…if it’s possible to have actually for example our collaborators in the US help out with some of the procurement to cut out on the delays…the issues that we get are are really outside the system, either in the programme or in the procurement, and these are really outside the Unit. And maybe the other thing like I said if the procurement-if they can have a section for scholars. Because what we do-our work is really time bound. And delays in our programme actually gives us a lot of pressure, a lot of tension”* -- IDI Male 2, Ph.D.

### Resource and financial mobilization

Streamline consistency across all training projects to ensure adequate funding for research and stipends, enabling trainees to conduct their research effectively and access essential research materials, tools, and technologies, thereby facilitating the advancement of their projects.

> *“If possible all PHD students should have research funding and this is based from my own experience cause I did not have, though I got I am just looking at it if a program for example says we give you a stipend, we pay you school fees…look for research funding…”* -- IDI Male 3, Ph.D.

## DISCUSSION

The results of our study provide insight into the multifaceted experiences and perceptions of alumni and continuing scholars affiliated with the Capacity Building Unit (CBU) at IDI. Amidst the challenges, such as procurement delays, supervisory issues, and limited research funds, a resounding affirmation of the invaluable support provided by the CBU in nurturing trainee research and career development journeys emerges. From personalized mentorship to immersive learning experiences, peer collaboration platforms, and grant support, the CBU has emerged as a beacon of support for scholars navigating the complex landscape of academic and research endeavors (17). Our sustainable capacity-building model at IDI is based on peer-to-peer and senior-to-junior mentorship approaches. Our trainees highlighted individual mentorship and supervision, training, and learning opportunities through short courses, statistical and publication support, research dissemination and evaluation meetings, grants application support, access to institutional resources, networking with renowned research experts, and additional funding for their research projects as benefits of belonging to the program. We see a similar model by Balandya et al., which utilizes the same vertical and horizontal mentoring strategies, creating a mentored hierarchical pyramid crucial for the succession of research expertise. Similarly, trainees reported having achieved substantial milestones, including publications, participation in research short course training, enrolment in Ph.D programs, involvement in ongoing research projects, and grant applications (18).

Our findings align with the sentiments of previous research, which emphasizes the pivotal role of mentorship, access to resources, and institutional backing in fostering the growth and success of scholars (19, 20). Our study extends the discourse by uncovering nuanced differences in perceptions between alumni and continuing scholars. While alumni reminisce fondly about their transformative experiences at CBU, continuing scholars offer constructive feedback, highlighting areas for improvement and refinement within the program. This disparity underscores the dynamic nature of capacity-building initiatives and the imperative of ongoing evaluation and adaptation to meet evolving needs (19).

Despite the benefits mentioned, several challenges scholars encounter include inadequate research funding and stipends, procurement delays, supervisory delays, and COVID-19 interruptions, similar to those reported in other studies. The COVID-19 disruptions led to delays in data collection, slow and expensive procurement of research supplies, and fewer physical meetings with the research team due to the social distancing measures implemented as preventive measures during the pandemic. Our findings indicate a general reduction in scholar-related challenges faced by continuing trainees and alumni during and after their tenure at the CBU compared to before joining the Unit. Nevertheless, a notable number of respondents reported challenges (21), particularly during the pre- and in-training phases of training under the CBU. This indicates that while the CBU may have mitigated some challenges through streamlining the mentorship structure, providing relevant training opportunities, and supporting economic infrastructure, there are still areas, such as additional research funding, procurement, and delays in supervisory feedback, where improvement may be needed to support scholars throughout their academic journey. The study’s findings highlight the pivotal role of mentorship, access to resources, and institutional support in fostering the growth and success of scholars. A supportive environment that facilitates conducive learning, collaboration, and innovation ecosystem, such as the IDI CBU, plays a critical role in shaping the trajectory of research scholars’ careers and catalyzing nuances in scientific advancement.

Among the institutional recommendations suggested were surveys to identify barriers to career growth, allowing mentees to choose their own mentors, identifying and supporting promising students from medical schools, improving procurement processes, offering short-term fellowships at the Institute, providing adequate research funding and stipends, and strengthening peer group leadership and enrollment. This is consistent with findings from other settings in Tanzania, where a unifying model for research mentorship comprised of senior researchers, early and middle-career students in higher institutions highlighted the need for an enabling environment to foster collaborations, resource mobilization, training, career growth, and mentorship as the basic requirement for sustainable research outputs at institutions for higher learning (9).

### Strength

This study employs a mixed-methods approach, combining quantitative and qualitative results to triangulate the study’s findings. It further paves the way for future research avenues to deepen our understanding of capacity-building initiatives and their enduring impact on scholars’ trajectories. Longitudinal investigations tracking participants over extended periods could offer insights into the sustained efficacy of the CBU and illuminate the factors contributing to long-term success.

### Limitation

The reliance on self-reported data introduces the possibility of response bias, necessitating cautious interpretation of the findings.

### Conclusion

Our findings underscore the importance of sustained investment in research capacity-building programs and highlight the enduring impact of mentorship, collaboration, and institutional support in fostering scientific excellence and innovation. The study findings will be utilized as a benchmark for continuous quality improvement of the capacity-building process at our Institution.

## Data Availability Statement

All data generated or analyzed during this study are included in this published article and/supplementary material. Further inquiries can be directed to the corresponding author.

## Ethical Consideration

The study was approved by The AIDS Support Organisation Research Ethics Committee (accreditation number: UG-REC-009). The study also received approval from the UNCST (study reference number: HS1100ES). Informed consent was obtained from each participant before their responses to the questionnaire and interviews. Administrative clearance was obtained from IDI to carry out this study.

## Authors’ contributions

AN conceptualized and wrote the protocol, supported study administration and coordination, including obtaining all regulatory and administrative approvals, and participated in data collection, analysis, and drafting the initial and final manuscript. BC contributed to the conception, design, and review of the final version, serving as the senior author. AK contributed to the study design and review of the protocol. VN and JM contributed to the study design, data analysis, and interpretation of quantitative data, as well as the review of the final manuscript. AT contributed to the design, analysis, and interpretation of the qualitative data and reviewed the final manuscript. CSW and ML contributed to the design and review of the final manuscript. YCM reviewed the final manuscript. All authors read and approved the final manuscript.

## Acknowledgments

We extend our appreciation to the CBU scholars for their participation in the study. We thank our research assistants, Ashaba Andrew, Nicolate Nekesa, Monica Agena, and Denis Mayambala, for their efforts during the implementation of our study. We sincerely appreciate the support rendered by the research department leadership and office at the Infectious Diseases Institute.

